# CSF Aβ38 levels are associated with Alzheimer-related decline: implications for γ-secretase modulators

**DOI:** 10.1101/2021.01.31.21250702

**Authors:** Nicholas C. Cullen, Shorena Janelidze, Sebastian Palmqvist, Erik Stomrud, Niklas Mattsson-Carlgren, Oskar Hansson

## Abstract

**Objective:** Shorter Aβ species might modulate disease progression in Alzheimer’s disease (AD). Here we studied whether Aβ38 levels in cerebrospinal fluid (CSF) are associated with risk of developing AD dementia and cognitive decline.

**Methods:** CSF Aβ38 levels were measured in 656 individuals across two clinical cohorts – the Swedish BioFINDER study and the Alzheimer’s Disease Neuroimaging Initiative (ADNI). Cox regression models were used to evaluate the association between baseline Aβ38 levels and risk of AD dementia in AD-biomarker positive individuals (AD+; determined by CSF P-tau/Aβ42 ratio) with subjective cognitive decline (SCD) or mild cognitive impairment (MCI). Linear mixed effects models were used to evaluate the association between baseline Aβ38 levels and cognitive decline as measured by MMSE in AD+ participants with SCD, MCI or AD dementia.

**Results:** In the BioFINDER cohort, high Aβ38 levels were associated with slower decline in MMSE (β = 0.30 points / sd., P = 0.001) and with lower risk of conversion. To AD dementia (HR = 0.83 per sd., P = 0.03). In the ADNI cohort, higher Aβ38 levels were associated with less decline in MMSE (β = 0.27, P = 0.01), but not risk of conversion to AD dementia (P = 0.66). Aβ38 levels in both cohorts remained significantly associated with both outcomes when adjusted for CSF P-tau levels and remained associated with cognition when adjusted for CSF Aβ42 levels.

**Conclusions:** Higher CSF Aβ38 levels are associated with lower risk of AD-related changes in two independent clinical cohorts. These findings may have implications for γ-secretase modulators as potential disease-altering therapy.

## Introduction

Despite a number of promising clinical trials of Alzheimer’s disease (AD) there is no approved treatment to delay disease progression [1,2]. Currently, the most widely accepted explanation for AD pathogenesis is the amyloid cascade hypothesis, which proposes that AD is primarily initiated by accumulation of the beta-amyloid (Aβ) peptide into senile plaques, sequentially followed by the accumulation of misfolded tau protein into tangles, neuronal loss, and cognitive decline along with loss of independence in activities of daily living (ADL) [3].

An expected consequence of the amyloid cascade hypothesis is that modulating the production of Aβ levels in the brain should prevent downstream effects of this pathology and thereby slow the disease course. It is precisely this mechanism which has been the target of recent AD therapies, although none of these therapies have been shown to affect disease progression as measured by cognitive tests [4–7]. Resolving the disagreement between overwhelming evidence speaking for amyloid’s role as disease driver and the failure of anti-amyloid therapies is therefore a major open question in the AD research field.

One proposed explanation for this failure is that the canonical view of amyloid accumulation may be oversimplified, particularly as it relates to the 42-amino acid long peptide (Aβ42) which has been the primary focus of fluid biomarker studies. Increasing evidence suggests that the relative abundance of different Aβ isoforms, especially those shorter than Aβ42, may play a more decisive role in AD pathogenesis than previously thought [8,9]. For instance, many presenilin mutations known to cause a familial form of AD do not directly result in higher Aβ42 levels in the brain, but rather disturb the relationship between Aβ42 and shorter Aβ species through a loss-of-function mechanism [10–12]. A role for shorter Aβ species in AD development could explain why targeting of amyloid aggregates expressed primarily by Aβ42 levels is not sufficient to curb the trajectory of AD. Investigating the association between shorter Aβ peptides and AD-related changes is therefore important for understanding amyloid accumulation, particularly as it relates to disease-altering therapies.

In the present study, we took a clinical approach to this question by measuring Aβ38 levels in cerebrospinal fluid (CSF) and characterizing them with regards to risk of developing AD dementia as well as cognitive decline. Our analysis was performed in two large, independent cohorts comprised of individuals spread broadly across the AD spectrum. Our primary aim was to understand whether CSF Aβ38 levels relate to AD-relevant outcomes and thereby shed more light on the complex relationship between the amyloid protein and AD.

## Methods

### Study design and participants

Participants recruited for the Swedish BioFINDER study were enrolled consecutively between 2010 and 2014 (clinical trial no. NCT03174938). Individuals with mild cognitive impairment (MCI) were recruited consecutively and were thoroughly assessed by physicians with special competence in dementia disorders. The inclusion criteria were the following: referred to a memory clinic due to possible cognitive impairment, not fulfilling the criteria for dementia, MMSE score 24–30, at least 60 years old, and fluent in Swedish [13]. AD individuals fulfilled the NIA-AA criteria for probable AD [14]. A diagnosis of subject cognitive decline (SCD) was done in coordination with an experienced neurologist. All participants were enrolled consecutively after being referred to a memory clinic. All relevant ethical committees approved the BioFINDER study and all study participants gave written informed consent.

Additional data was analyzed from participants in the Alzheimer’s Disease Neuroimaging Initiative (ADNI) study, which was launched in 2003 as a public-private partnership. Participants in the ADNI study have been recruited from more than 50 locations across the United States and Canada. Inclusion and exclusion criteria for ADNI have been described in detail previously [15]. Briefly, all ADNI participants were between the ages of 55 and 90 years, had completed at least six years of education, were fluent in Spanish or English, and had no significant neurologic disease other than AD. Regional ethical committees of all institutions approved the ADNI study and all study participants gave written informed consent. ADNI data was downloaded from http://adni.loni.usc.edu on 2020/03/01.

The present analysis included only AD-biomarker positive (“biomarker-positive”) participants, determined based on an abnormal CSF P-tau/Aβ42 ratio for which cutoffs have been previously established in both cohorts [16].

### Predictors and outcomes

Demographic characteristics including age, sex, and education were collected from all participants. Various biomarkers of amyloid processing or pathology – Aβ38, Aβ40, Aβ42, and amyloid precursor protein (sAPP; available in BioFINDER only) – along with tau phosphorylated at threonine 181 (P-tau) were measured in CSF at baseline in all participants. All biomarkers were natural log-transformed prior to analysis in order to obtain a more normal distribution of biomarker values. In BioFINDER, Aβ38, Aβ40, Aβ42, P-tau, and sAPP levels were measured using a standard ELISA assay (Euroimmun, Lubeck, Germany). In ADNI, Aβ38, Aβ40, and Aβ42 levels in CSF were measured using a 2D-UPLC tandem mass spectrometry method at the University of Pennsylvania (first made publicly available in February 2020). CSF P-tau levels in ADNI were measured using the Elecsys platform (Roche, Basel Switzerland).

The primary cognitive outcome was longitudinal change in cognition as measured by the Mini-Mental State Examination (MMSE) scale. MMSE is a cognitive test which is highly relevant to cognitive changes in AD and is often used as a basis for making a clinical diagnosis or inclusion into clinical trials [17]. The primary clinical outcome was conversion to AD dementia at any time during longitudinal follow-up. Clinical status was evaluated and recorded at each follow-up visit by a physician experienced in dementia disorders.

### Statistical Analysis

Linear mixed effects (LME) modelling was used to assess the relationship between continuous Aβ38 levels (adjusted for age, sex, and education) and longitudinal change in MMSE. Additional LME models were fit which also included covariate adjustment for CSF Aβ42 or P-tau levels. LME models had random intercepts and slopes with an interaction term between time and Aβ38 levels and an interaction term between time and Aβ42 or P-tau levels for models which also included those biomarkers.

Cox regression modelling was used to assess the association between continuous Aβ38 levels (adjusted for age, sex, and education) and conversion to AD dementia during longitudinal follow-up. Additional cox regression models were fit which also included covariate adjustment for either CSF Aβ42 or P-tau levels. All participants were right-censored (i.e., the last follow-up visit was considered as either the latest visit if the participant was never diagnosed with AD dementia or the visit when diagnosis of AD dementia occurred) and the proportionality of hazards assumption was assessed using Schoenfeld residuals.

All code was written in the R programming language (v4.0.0) and all significance tests were two-sided with alpha = 0.05 as significance threshold.

## Results

### Cohort characteristics

In the BioFINDER cohort (Table 1), we included 338 biomarker-positive participants classified as either SCD (n = 54), MCI (n = 150), or AD (n = 134). The average age was 72.5 ± 6.8 years, 52.1% of participants were female, and average education was 11.0 ± 3.5 years. In the ADNI cohort (Table 2), we included 318 biomarker-positive participants classified as either SCD (n = 17), MCI (n = 192), or AD (n = 109). The average age was 73.1 ± 7.4 years, 54.7% of participants were female, and average education was 15.9 ± 2.8 years. In testing the relationship between biomarkers in both cohorts (Figure 1), we found that CSF Aβ38 was significantly correlated with Aβ42 (r = 0.44, P < 0.0001 in BioFINDER; r = 0.54, P < 0.0001 in ADNI) and with P-tau (r = 0.37, P < 0.0001 in BioFINDER; r = 0.53, P < 0.0001 in ADNI).

**Table 1:**
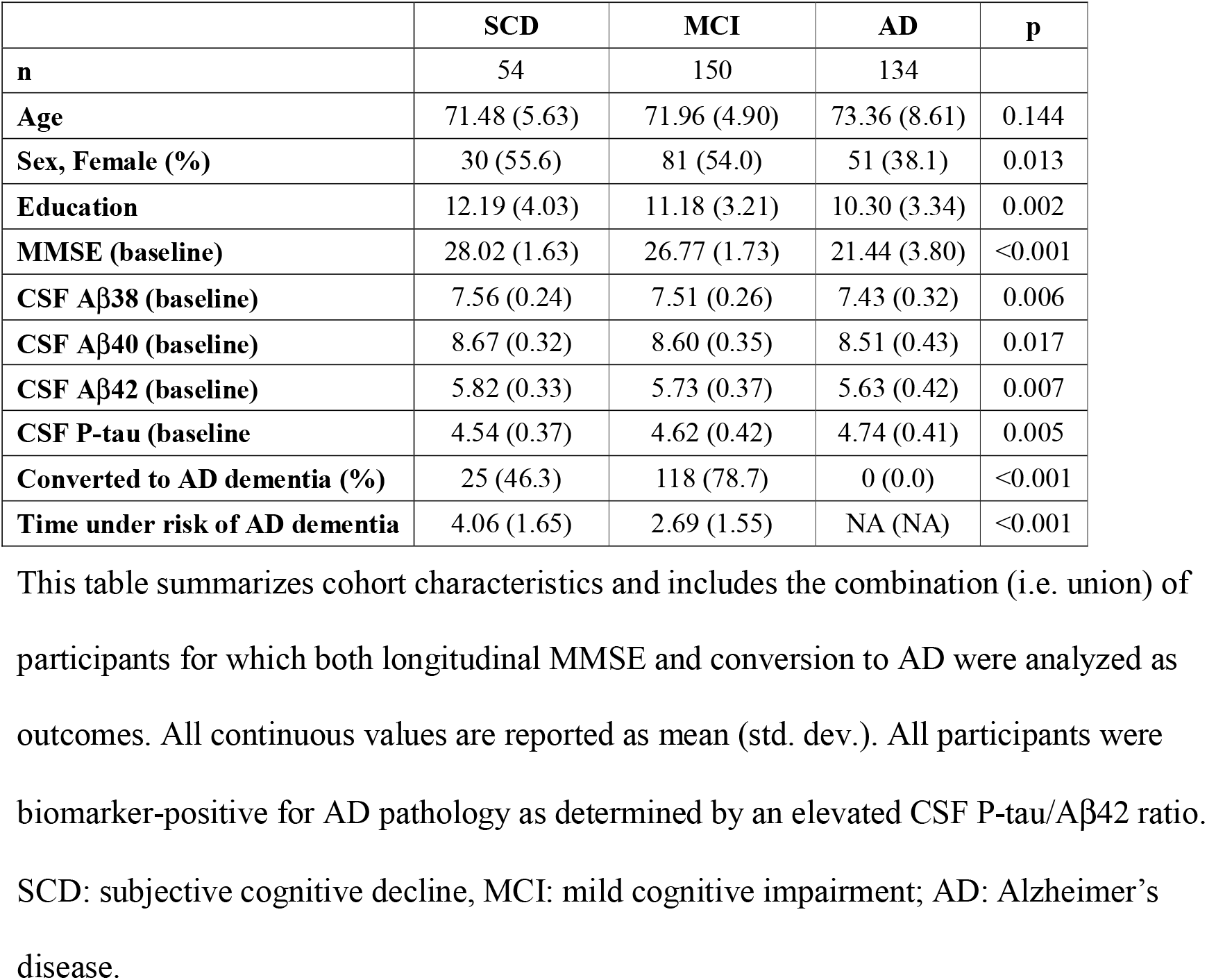
Cohort characteristics – BioFINDER.

**Table 2:** Cohort characteristics – ADNI.

|  | **SCD** | **MCI** | **AD** | **p** |
| --- | --- | --- | --- | --- |
| **n** | 17 | 192 | 109 |  |
| **Age** | 73.96 (5.25) | 72.56 (6.95) | 73.94 (8.34) | 0.263 |
| **Sex, Female (%)** | 13 (76.5) | 81 (42.2) | 50 (45.9) | 0.024 |
| **Education** | 16.65 (2.67) | 15.95 (2.76) | 15.55 (2.72) | 0.224 |
| **MMSE (baseline)** | 29.29 (0.85) | 27.45 (1.82) | 23.04 (2.03) | <0.001 |
| **CSF Aβ38 (baseline)** | 7.58 (0.30) | 7.56 (0.30) | 7.42 (0.33) | 0.001 |
| **CSF Aβ40 (baseline)** | 9.03 (0.28) | 9.02 (0.28) | 8.90 (0.32) | 0.005 |
| **CSF Aβ42 (baseline)** | 6.66 (0.31) | 6.56 (0.32) | 6.48 (0.35) | 0.042 |
| **CSF P-tau (baseline)** | 3.50 (0.35) | 3.53 (0.38) | 3.61 (0.38) | 0.164 |
| **Converted to AD dementia (%)** | 0 (0.0) | 102 (53.1) | 0 (0.0) | <0.001 |
| **Time under risk of AD dementia** | 3.50 (1.88) | 2.84 (2.11) | NA (NA) | 0.213 |

This table summarizes cohort characteristics and includes the combination (i.e. union) of participants for which both longitudinal MMSE and conversion to AD were analyzed as outcomes. All participants were biomarker-positive for AD pathology as determined by an elevated CSF P-tau/Aβ42 ratio. SCD: subjective cognitive decline, MCI: mild cognitive impairment; AD: Alzheimer's disease.

**Figure 1:**
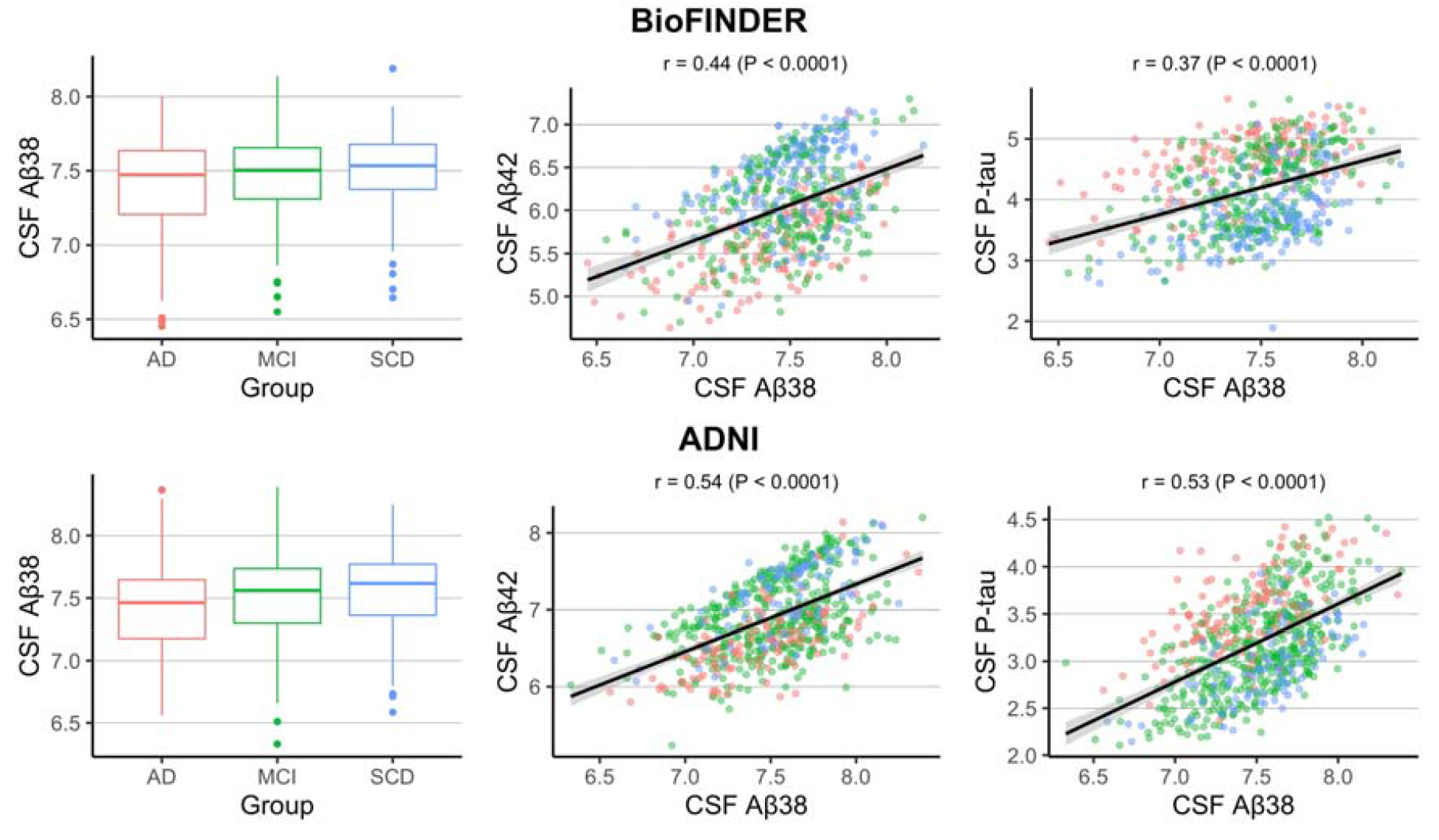
Distribution of CSF Aβ38 levels across diagnostic groups and cohorts and their association with CSF Aβ42 and CSF P-tau. This figure shows how CSF Aβ38 levels are distributed across diagnostic groups (left-most panels) and how CSF Aβ38 levels relate to CSF Aβ42 (center panels) and CSF P-tau (right-most panels) levels in the BioFINDER (top three panels) and ADNI (bottom three panels) cohorts. Association between biomarkers was tested using Pearson correlation. Datapoints in the scatter plot are colored according the same scheme in the boxplot (far left): red = AD, green = MCI, blue = SCD.

### Association with longitudinal decline in cognition

In the BioFINDER cohort, higher CSF Aβ38 levels adjusted only for demographics (age, sex, and education) were associated with less decline in MMSE over time (β = 0.30 points / year per std. of biomarker change, P = 0.001). Higher Aβ38 levels adjusted for Aβ42 levels were also associated with less decline in MMSE over time (β = 0.25, P = 0.03); Aβ42 in the same model was not significantly associated with MMSE change (P = 0.57). Higher Aβ38 levels with additional adjustment for P-tau were associated with less decline in MMSE over time (β = 0.76, P < 0.0001); P-tau in the same model was also significantly associated with MMSE change (β = −0.94, P < 0.0001). These results are displayed graphically in Figure 2.

**Figure 2:**
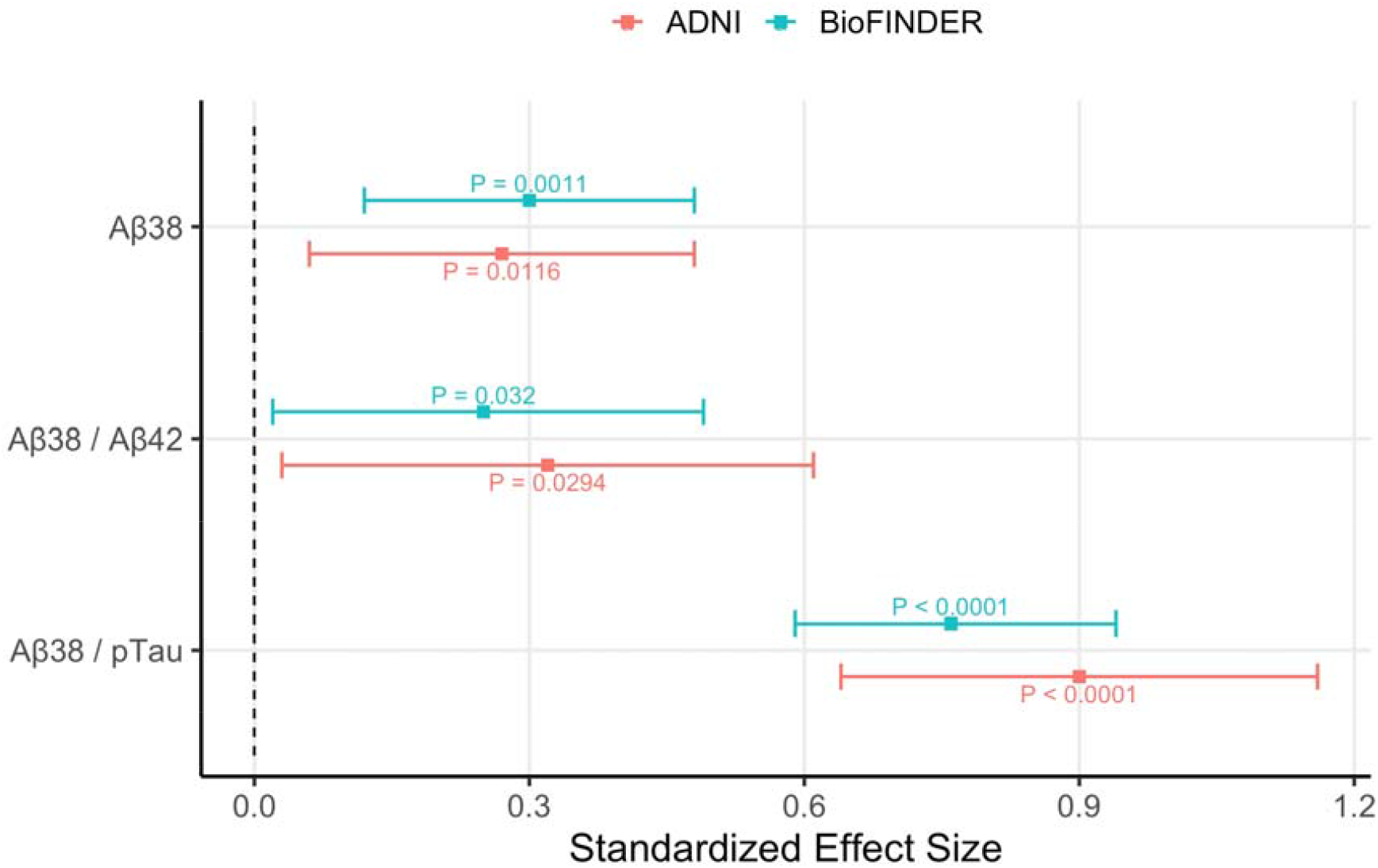
Association between CSF Aβ38 and longitudinal cognition across cohorts. This figure displays results from linear mixed effects analysis in the BioFINDER and ADNI cohorts to investigate the association between longitudinal MMSE and Aβ38 alone, Aβ38 adjusted for Aβ42, and Aβ38 adjusted for P-tau levels. All models were additionally adjusted for age, sex, and education. Coefficients are displayed both for the effect of Aβ38 on baseline MMSE (“baseline” in the figure) and on change in MMSE over time (“slope” in the figure).

In the ADNI cohort, higher Aβ38 levels adjusted only for demographics (age, sex, and education) were associated with less decline in MMSE over time (β = 0.27, P = 0.01). Higher Aβ38 levels with additional adjustment for Aβ42 were also associated with less decline in MMSE over time (β = 0.32, P = 0.03); Aβ42 in the same model was not significantly associated with MMSE change (P = 0.61). Finally, higher Aβ38 levels with additional adjustment for P-tau were associated with less decline in MMSE over time (β = 0.90, P < 0.0001); P-tau in the same model was also significantly associated with MMSE change (β = −0.95, P < 0.0001). These results are displayed graphically in Figure 2.

### Association with risk of developing AD dementia

In the BioFINDER cohort, higher Aβ38 levels adjusted only for demographics were associated with lower risk of AD dementia (HR = 0.83 higher odds per std. of biomarker change, 95% CI [0.71, 0.98], P = 0.03), while higher Aβ38 levels additionally adjusted for Aβ42 trended towards an association with lower risk of conversion (HR = 0.85 [0.69, 1.05], P = 0.12), and higher Aβ38 levels additionally adjusted for P-tau were strongly associated with lower risk of conversion (HR = 0.56 [0.46, 0.69], P < 0.0001). These results are displayed graphically in Figure 3.

**Figure 3:**
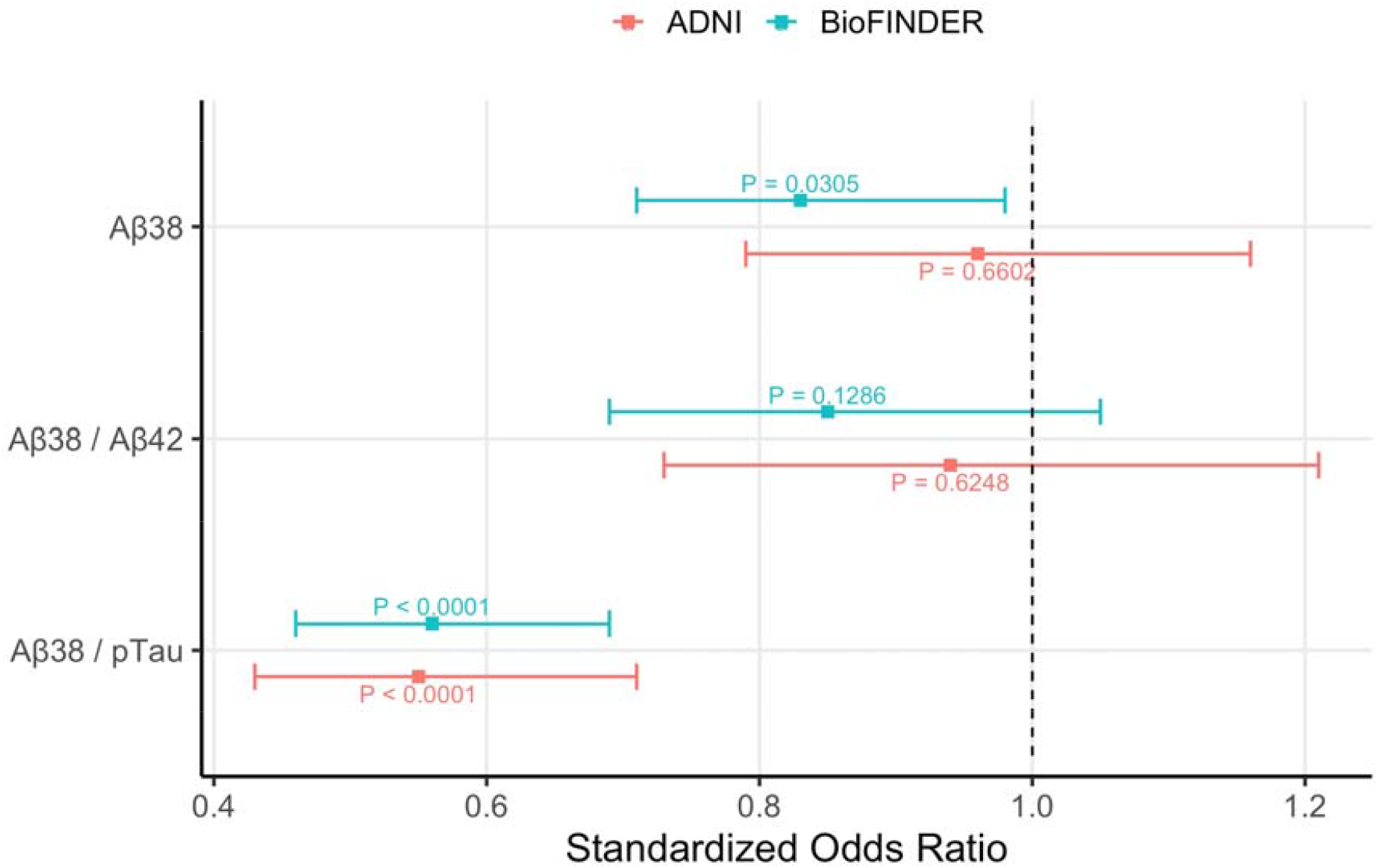
Association between CSF Aβ38 and clinical conversion across cohorts. This figure displays results from Cox regression analysis in the BioFINDER and ADNI cohorts to investigate the association between risk of developing AD dementia and Aβ38 alone, Aβ38 adjusted for Aβ42, and Aβ38 adjusted for P-tau levels. All models were additionally adjusted for age, sex, and education. Coefficients represents the change in odds of converting to AD dementia for each standard deviation increase in Aβ38 levels.

In the ADNI cohort, there was not a significant association with conversion to AD dementia when Aβ38 levels were adjusted only for demographics (HR = 0.96 [0.79, 1.16], P = 0.66), and there was not a significant association when Aβ38 was adjusted for demographics and Aβ42 (HR = 0.94 [0.73, 1.21], P = 0.62). Still, higher Aβ38 levels adjusted for demographics and P-tau were strongly associated with lower risk of conversion to AD dementia (HR = 0.55 [0.43, 0.71], P < 0.0001). These results are displayed graphically in Figure 3.

#### Analysis of other Aβ biomarkers

We performed the same analyses above and in the same groups but using CSF Aβ40 as the variable of interest instead of CSF Aβ38. In BioFINDER, higher CSF Aβ40 levels adjusted only for demographics were associated with less decline in MMSE (β = 0.30 points / year per std. of biomarker change, P = 0.01; same effect size as for Aβ38). Aβ40 levels were also associated with less decline in MMSE when adjusted additionally for CSF Aβ42 (β = 0.27, P = 0.04; larger than for Aβ38) and CSF P-tau (β = 0.72, P < 0.0001; smaller than for Aβ38). In ADNI, higher Aβ40 levels adjusted for only demographics were not associated with higher change in MMSE (β = 0.21, P = 0.051), nor when also adjusted for CSF Aβ42 (β = −0.09, P = 0.15), but were significant when adjusted for CSF P-tau (β = 0.84, P < 0.0001). To note, the standardized effect sizes for Aβ40 in the ADNI cohort were smaller in magnitude for all models than those that instead included Aβ38.

With regards to conversion to AD dementia in the BioFINDER cohort, CSF Aβ40 levels adjusted only for demographics were weakly associated with conversion to AD dementia (HR = 0.84 lower odds per std. of biomarker change, P = 0.048). As with Aβ38, higher Aβ40 levels were not associated with conversion to AD dementia when adjusted further for Aβ42 (HR = 0.86, P = 0.21), but did have a significant association when adjusted further for P-tau (HR = 0.61, P < 0.0001). The standardized effect size for Aβ40 was smaller for all modes in the BioFINDER cohort compared to the same models with Aβ38. In ADNI,

CSF Aβ40 levels were not associated with conversion to AD dementia when adjusted only for demographics (HR = 1.03, P = 0.78) or additionally for Aβ42 (HR = 1.08, P = 0.61), but were significantly associated when adjusted further for P-tau (HR = 0.60, P = 0.0003). The standardized effect size for significant Aβ40 models in the ADNI cohort was again smaller than the corresponding models that included Aβ38 instead.

A similar analysis of CSF sAPP levels (available only in BioFINDER) showed no association with change in MMSE (β = −0.01, P = 0.94) or conversion to AD dementia (HR = 0.94, P = 0.65).

## Discussion

There is a great need in the AD research field to explain why overwhelming evidence points to amyloid being the key driver of AD pathogenesis while anti-amyloid therapies have mostly failed to affect disease progression in late-stage clinical trials. One proposed explanation to recent trial failures has been to question inherent factors of the trials themselves – e.g. inclusion criteria, choice of endpoint, too late treatment initiation, or statistical power [18].

However, more recent AD trials have included stringent biomarker inclusion criteria, sophisticated composite clinical endpoints, and large numbers of participants [5,6,19].

Assuming then that these trial failures are due to biological factors, another response has been to instead question the entire amyloid cascade hypothesis by suggesting that amyloid accumulation may be an indirect effect rather than primary cause of AD [20]. However, existence of early-onset familial AD caused by mutations in the *APP, PSEN1*, and *PSEN2* genes – all part of the Aβ processing machinery – suggests that the true explanation for failure of anti-amyloid therapies should still retain the integrity of the amyloid cascade hypothesis [21]. One possible mechanism could be that there is a more complex interaction between different Aβ peptides than previously appreciated, which would explain why targeting of Aβ42 alone may not be sufficient to alter disease progression. Recent evidence suggests that lower levels of shorter Aβ peptide levels or lower ratio of shorter to longer Aβ peptides could be an important factor in Aβ toxicity [9,22,23].

Our results in the current study support this hypothesis from a clinical perspective, as we demonstrated that higher CSF Aβ38 levels are associated with less cognitive decline and lower risk of developing AD dementia in individuals who are biomarker-positive (see Table 3 for summary of evidence). Our finding that higher CSF Aβ38 levels are protective even in the presence of significant AD pathology in the brain (we only included biomarker-positive individuals) should motivate further studies to understand the molecular underpinning of a potential protective mechanism from Aβ38, and possibly even Aβ40. For instance, it is unclear if Aβ38 levels modulate the development of tau pathology in individuals who already reached thresholds for Aβ positivity pathology.

**Table 3:** Qualitative summary of results.

| Outcome | Aβ38 | | Aβ38 (+ Aβ42) | | Aβ38 (+ P-tau) | |
| --- | --- | --- | --- | --- | --- | --- |
|  | BioFINDER | ADNI | BioFINDER | ADNI | BioFINDER | ADNI |
| Baseline MMSE | *** | *** | * | * | *** | *** |
| Change in MMSE | *** | * | * | * | *** | *** |
| Conversion to AD | * | NS | NS | NS | *** | *** |

This table summarizes evidence for the association of Aβ38 levels with AD-related changes in the BioFINDER and ADNI cohorts. Asterisks in the table represent a significant association between the biomarker model (top row) and the outcome (left-most column) in the given cohort (second-to-top row) based on tests from linear mixed effects modelling (for baseline MMSE and change in MMSE) or Cox regression modelling (for conversion to AD). All models were adjusted for age, sex, and education. NS = Not Significant; \* = $P < 0.05$ ; \*\* = $P < 0.005$ ; \*\*\* = $P < 0.0005$ .

The validation of our findings in two independent cohorts with differing demographic profiles – ADNI participants have high educational attainment on average and are primarily typical amnestic AD cases, while the BioFINDER cohort is more heterogenous in demographical and diagnostic makeup – adds validity to our results. Importantly, the effect sizes for Aβ38 in the statistical models were generally stronger than those for Aβ40 or sAPP – indicating a specific effect of Aβ38 rather than simply an effect of total Aβ production or APP cleavage. Taken together, then, it is unlikely that our findings could be due to systematic changes related to CSF collection, volume, or measurement.

These findings are of particular importance due to the renewed interest in γ-secretase modulators (GSMs), a class of drugs which reduces Aβ42 production while maintaining total Aβ production by blocking cleavage of amyloid precursor protein (APP) at specific γ-secretase cleavage sites [24,25]. In comparison to previously tested γ-secretase inhibitors (GSIs), which had untenable off-target effects in past clinical trials, GSMs do not alter total Aβ production and thus do not compromise the broader biological role of γ-secretase [25,26]. Previous studies of GSMs have shown that the Aβ38 peptide does not exhibit any toxicity *in vivo* (nor does it accumulate into plaques following overexpression in mice) and can even protect against Aβ42-associated dysfunction [22]. However, while the Aβ42/Aβ40 ratio has been widely implicated in both clinical studies of AD and animal studies of GSMs (primarily as a proxy for brain Aβ build-up), few clinical studies prior to ours have investigated Aβ38 levels [27–29].

To summarize, our results suggest that further investigations should be undertaken to understand whether increasing the relative levels of shorter Aβ peptides such as Aβ38 is in fact an effective strategy to treat AD [30,31]. We provided clinical evidence here that higher Aβ38 levels are in fact associated with lower risk of AD-related changes, which may support the use of GSMs as an approach to altering AD progression. However, our understanding of the interaction between the different Aβ peptides is still lacking.

## Data Availability

Data is available upon request.

## Author contributions

NC, NMC and OH designed the study. NC performed statistical analyses and wrote the draft of the paper. SJ performed biomarker measurements. ES and SP managed clinical evaluations and logistics. NMC and OH supervised the work. All authors reviewed the paper for intellectual content.

## Acknowledgments

Work at the authors’ lab is funded by Knut and Alice Wallenberg Foundation, the Medical Faculty at Lund University, Region Skåne, the European Research Council, the Swedish Research Council, the Strategic Research Area MultiPark at Lund University, the Swedish Alzheimer Foundation, the Swedish Brain Foundation, The Parkinson foundation of Sweden, The Parkinson Research Foundation, the SkaCne University Hospital Foundation, the Swedish federal government under the ALF agreement, the Konung Gustaf V:s och Drottning Victorias Frimurarestiftelse, and the Bundy Academy.

